# A Cross-Sectional Analysis of Syncytiotrophoblast Membrane Extracellular Vesicles Derived Transcriptomic Biomarkers in Preeclampsia

**DOI:** 10.1101/2023.07.28.23293326

**Authors:** Toluwalase Awoyemi, Wei Zhang, Maryam Rahbar, Adam Cribbs, Prasanna Logenthiran, Shuhan Jiang, Gavin Collett, Ana Sofia Cerdeira, Manu Vatish

## Abstract

**Background:** Preeclampsia (PE) is a pregnancy-specific hypertensive disorder affecting 2-8% of pregnancies worldwide. Biomarker(s) for PE exists, but while these have excellent negative predictive value, their positive predictive value is poor. Extracellular vesicles released by the placenta into the maternal circulation, syncytiotrophoblast membrane extracellular vesicles - STB-EVs-have been identified as being involved in PE with the potential to act as liquid biopsies.

**Objective:** To identify differences in STB-EV and placenta transcriptome between PE and normal pregnancy (NP).

**Methods:** We performed RNA-sequencing (RNA-seq) on placental tissue, medium/large and small STB-EVs from PE (n=6) and NP (n=6), followed by bioinformatic analysis to identify targets that could be used in the future for EV-based diagnostic tests for preeclampsia. Some of the identified biomarkers were validated with real-time polymerase chain reactions.

**Results:** Our analysis identified and verified the differential expression of FLNB, COL17A1, SLC45A4, LEP, HTRA4, PAPP-A2, EBI3, HSD17B1, FSTL3, INHBA, SIGLEC6, and CGB3. Our analysis also identified interesting mechanistic processes via an in-silico prediction of STB-EV-based mechanistic pathways.

**Conclusions:** In this study, we identified potential biomarkers and mechanistic gene pathways that may be important in the pathophysiology of PE and could be further explored in future studies.

**Funding:** This research was funded by the Medical Research Council (MRC Programme Grant (MR/J0033601) and the Medical & Life Sciences translational fund (BRR00142 HE01.01)

## Introduction

Preeclampsia (PE) is a multisystemic hypertensive disorder that affects approximately 2-8% of pregnancies worldwide (Lisonkova & Joseph, 2013). It is a pregnancy-specific complication which results in hypertension (systolic blood pressure ≥ 140mmHg / diastolic pressure ≥ 90mmHg) and proteinuria (protein/creatinine ratio of ≥ 30 mg/mmol or more), or evidence of maternal acute kidney injury, liver dysfunction, neurological abnormalities, hemolysis, thrombocytopenia, and/or fetal growth restriction and in severe cases, death(Brown et al., 2018). Preeclampsia in particular, early onset preeclampsia (EOPE) has been referred to as a two-stage process, starting with mal-placentation which results in syncytiotrophoblast stress (stage 1) and ending in maternal end-organ damage (stage 2)(C. W. G. Redman et al., 2022). However, the direct cause of PE (i.e., mechanisms of placental dysfunction) is still under study, with the only available treatment being the delivery of the placenta (C. Redman, 2014).

Syncytiotrophoblast membrane extracellular vesicles (STB-EVs) are lipid bilayer spherical structures of placental origin which can be classified based on their size and biogenesis pathways into medium/large and small STB-EVs (Yáñez-Mó et al., 2015). Small STB-EVs are less than or equal to 200 nm in size and formed through multivesicular bodies, whereas medium/large EVs are between 201 nm to 1000 nm and released directly through the budding of the plasma membrane (Théry et al., 2018). Their cargo consists of surface proteins, encapsulated proteins, and different subclasses of RNAs such as long noncoding RNAs, messenger RNAs, transfer RNAs, and micro RNAs (Colombo et al., 2014), which may be released into target cells through fusion of STB-EVs with distant cells. This ability of STB-EVs further implicates them as potential pathogenic factors in PE. However, the transcriptome of the subtypes of STB-EVs has not been thoroughly explored. Previous studies have identified differentially expressed genes (DEGs) from PE placental tissue such as *Sialic acid-binding Ig-like lectin 6 (SIGLEC6), Vascular endothelial growth factor receptor 1(VEGFR1), Adrenomedullin (ADM),* and *Pappalysin-2 (PAPP-A2)* (Rumer et al., 2013; Tsai et al., 2011a), *Basic Helix-Loop-Helix Family Member E40 (BHLHE40), Divergent-Paired Related Homeobox (DPRX),* and *HtrA Serine Peptidase 4 (HTRA4)* (Ren et al., 2021) compared to normal placental tissue. However, none to the best of our knowledge have explored this in the context of STB-EVs between PE and normal pregnancy (NP).

We hypothesized that 1) the transcriptome of the placental tissue and STB-EVs are different between PE and NP and 2) analyzing these different sample sub-types would allow for more comprehensive and holistic profiling. We believe this strategy increases the probability of detecting relevant biomarkers and mechanistic pathways in PE. We performed RNA-sequencing (RNA-seq) on placental tissue, medium/large and small STB-EVs from PE and NP. Our analysis revealed novel biomarkers and new insights into possible mechanisms of preeclampsia. This knowledge may help to inform future extracellular vesicle-based diagnostic tests, mechanistic experiments and ultimately, the development of new therapies.

## Methods

### Ethics approval and patient information

We obtained ethical approval from the Central Oxfordshire Research Ethics Committee C (REFS 07/H0607/74 & 07/H0606/148). We obtained written informed consent from pregnant women undergoing elective caesarean sections before labour onset at the Women’s Centre, John Radcliffe Hospital, Oxford. Placentas from normal (NP) n=12 and preeclamptic (PE) n=12 pregnancies were collected and perfused within ten minutes of delivery. We defined NP as singleton pregnancy with no history of preeclampsia, hypertensive disorders, or other complications in pregnancy. Patients with preeclampsia were defined as the co-occurrence of *de novo* hypertension (blood pressure > 140/90 mmHg) and proteinuria (protein/creatinine ratio ≥ 30 mg/mmol) and/or evidence of maternal acute kidney injury, liver dysfunction, neurological abnormalities, hemolysis, thrombocytopenia, and/or fetal growth restriction after week twenty of gestation according to the criteria of the International Society for the Study of Hypertension in Pregnancy(Brown et al., 2018). All PE patients used in this study were of early onset PE (diagnosed before 34 weeks gestation).

### Enrichment of STB-EVs by placental dual-lobe perfusion and serial ultracentrifugation

We have previously published our protocol of STB-EVs isolation through *ex vivo* dual lobe placental perfusion (Dragovic et al., 2015). Briefly, we identified a suitable cotyledon (devoid of calcifications, ischemia, or rupture) and cannulated a placental artery and vein perfusing the placenta for three hours at a 4-5 ml/min flow rate to obtain placenta perfusate. The placenta perfusate was centrifuged twice at 1,500 g for ten minutes at 4°C (*Beckman Coulter Avanti J-20XP centrifuge using a Beckman Coulter JS-5.3 swing-out rotor*) to remove cell debris. The supernatant was carefully pooled and spun at 10,000 g (10K) in a swing bucket centrifuge (*Beckman L80 ultracentrifuge and Sorvall TST28.39 swing-out rotor*) at 4^0^C for 30 minutes. The 10K STB-EV pellet was washed with filtered phosphate buffer saline (fPBS) followed by resuspension of the 10K STB-EV pellets in fPBS. An aliquot of the resuspended pellets was analysed to identify and characterise STB-EVs, while the rest were aliquoted to obtain a protein concentration around 2-5 µg/µl (measured using a Pierce bicinchoninic acid (BCA) protein assay) and immediately stored at -80°C. The post-10K supernatant was filtered through a 0.22 µm Millipore stericup filtration device, then spun at 150,000 g for 2 hours (*Beckman L80 ultracentrifuge with a Sorvall TST28.39 swing-out rotor*) and the 150K STB-EV pellets were washed, resuspended in fPBS and aliquoted like the 10K STB-EV pellets. This working stock was used for subsequent analysis. Our STB-EV enrichment and categorization process has been deposited on EV Track ([http://www.EVTRACK.org], **EV-TRACK ID: EV210382**) with a score of 78% correlating with excellent enrichment and categorization.

### Transmission electron microscopy

STB-EV pellets were diluted with fPBS to achieve an STB-EV solution with concentrations between 0.1-0.3 µg/µl. Ten microliter of the STB-EV pellet solution was applied to freshly glowing discharged carbon formvar 300 mesh copper grids for two minutes, blotted with filter paper, and stained with 2% uranyl acetate for ten seconds and air-dried. STB-EV pellets on the grid were negatively stained to enhance the contrast between STB-EVs pellets and the background. The grids were imaged using an FEI Tecnai 12 TEM at 120 kV with a Gatan OneView CMOS camera.

### Flow Cytometry

A BD LSRII flow cytometer (BD Biosciences) with a blue, violet, and red laser was used for all sample analyses. Daily quality control (QC) was run using CS&T beads (BD Biosciences). Photomultiplier tube (PMT) voltage determined by CS&T run was applied to all fluorescent detectors with exception for the side scatter (SSC) which was determined by Apogee Mix (1493, Apogee Flow System, UK). SSC PMT voltage that triggered 0.59 µm silica beads and above was applied to all 10K STB-EV pellets and analyzed. An SSC threshold of 200 was applied to remove background noise below 0.59 µm silica beads. A flow rate of 10 µl/min was achieved using the TruCount beads (BD science). For sample staining, 90 ml of 10K STB-EV pellet were incubated with ten ml of Fc receptor blocker (Miltenyl, UK) for 10 minutes at 4°C and then stained with phycoerythrin (PE) conjugated PLAP (for syncytiotrophoblast origin), PE Vio770 conjugated anti classical HLA class I and II (to exclude co-isolated non-placenta EVs and white blood cell (WBC) EV co-isolation), Pacific blue conjugated CD41 (to identify co-isolated platelet EVs) and CD235a (to identify co-isolated red blood cell (RBC) EVs) for ten minutes at room temperature in the dark. Stained samples were transferred to an Ultra free 0.2 µm filter unit (Millipore) and centrifuged at 800 g for three minutes to remove unbound antibodies and EVs smaller than the filter pore size. Ninety microliters of fPBS were used to recover 10K STB-EVs retained on the filter membrane. Recovered 10K STB-EVs were further stained with BODIPY FL N-(2-aminoethyl)-maleimide [505/513 nm] (Molecular Probes) at a final concentration of 0.5 nM in the dark at room temperature for ten minutes before samples were diluted to 500 ml and analyzed on the flow cytometer to check for events rate. When necessary, dilutions were made to achieve an events rate of ≤ 400 count/second and to reduce swarming. 10K STB-EV pellets were analyzed at 10 µl/minute for ten minutes and a total of 100 µl diluted samples was analyzed for each sample. Fluorescence minus one (FMO-1) for each fluorochrome and stained samples re-acquired after 2% Nonidet P-40 (NP-40) (Sigma) treatment were used as controls. Data and figures generated were generated with the Flowjo software version 10 (Tree Star Inc., Ashland, OR).

### Nanoparticle Tracking Analysis

We further characterised the 10K and 150K STB-EV pellets by nanoparticle tracking analysis [(NTA) NanoSight NS500 instrument equipped with a 405 nm laser (Malvern UK), sCMOS camera and NTA software version 2.3, Build 0033 (Malvern UK)]. Before sample analysis, instrument performance was checked with silica 100 nm microspheres (Polysciences, Inc.). The 10K and 150K STB-EV pellets were individually diluted in fPBS to a range of 1/100,000. The samples were automatically injected into the sample chamber with a 1 ml syringe with the following script used for EV measurements: prime, delay 5, capture 60, repeat 4. Images of the analyzed samples were captured on camera at level 12 (Camera shutter speed; 15 milliseconds and Camera gain; 350) and NTA post-acquisition settings were optimized and kept constant between samples. Each video recording was analyzed to infer STB-EVs size and concentration profile.

### Western Blot Analysis

We performed western blots on placental lysates (PL) and STB-EVs to further characterise and immune-phenotype. All STB-EVs pellets were probed with PLAP (for syncytiotrophoblast origin), CD63 and ALIX (to confirm the presence of extracellular vesicles), and Cytochrome C (as a negative EV marker) as recommended by the international society for extracellular vesicles (ISEV) (Théry et al., 2018). Following characterization and identification of extracellular vesicles in 10K and 150K STB-EV pellets, we renamed them to medium/large (m/l) and small (s) STB-EVs respectively.

### RNA-Sequencing Library Preparation and Sequencing

For sequencing, RNA extraction and sample preparations were performed for placenta tissue (discovery cohort-6 NP, 6 PE) with the RNeasy mini kit and STB-EVs (6 NP, 6 PE) with the miRCURY™ RNA isolation kit for biofluids (Exiqon Services, Denmark) based on manufacturer’s protocol. The samples were sent to the Wellcome Centre for Human Genetics (WCHG) for sequencing using standard Illumina protocol. Details can be found in the supplementary material.

### Bioinformatics and Statistical Analysis of Messenger RNA sequences

We performed bioinformatics analysis on Galaxy (https://usegalaxy.org/). We used FastQC (Galaxy Version 0.72+galaxy1) to obtain overall QC metrics and MultiQC (Galaxy Version 1.9+galaxy1) to amalgamate the QC metrics before and after trimming of adapters [trimmomatic (Galaxy Version 0.38.0)]. Alignment was referenced to reference Homo Sapiens build 38 (hg38) genomes obtained from Ensembl with *HISAT 2* (Galaxy Version 2.2.1+galaxy0). For each sample, *featureCounts* (Galaxy Version 2.0.1+galaxy1) was used to quantify genes based on reads that mapped to the provided hg38 genome. A count matrix was generated with the *Column Join on Collection* (Galaxy Version 0.0.3) tool. Differential expression analysis was done with the DESeq2 package (v.1.32.0 in R v.4.0.5). The reported p-values were adjusted for multiple testing using Benjamin Hochberg correction reported as false-discovery rate. An adjusted p-value less than 0.05 was taken as significant. Functional annotation was performed with ClusterProfiler (for Gene Ontology [GO] and KEGG) and Signalling pathway impact analysis (SPIA) with our transcriptome set as the background. The raw fastQ files and processed file have been deposited in NCBI and can be accessed with the following ID: GSE190973.

### Criteria for genes selection for validation

We chose a select group of DCGs between NP and PE to validate based on 1) fold change (genes with a high fold change [log FC ≥ (±)1] and adjusted P Value ≤10^-5^), 2) presence in at least two of the three sample types (placenta lysate, m/l STB-EVs, and sSTB-EVs), 3) placenta specificity or enrichment and, 4) previous molecules investigated by the group. Based on expression data at the protein and RNA level from the human protein atlas (Uhlen et al., 2017), genes are said to be placenta specific if they are expressed only in the placenta, while genes are placenta enriched if the placenta is one of the five with the highest expression.

### Quantitative polymerase chain reaction (qPCR) for mRNA validation

Following standard protocol, the high-capacity cDNA Reverse transcription kit (Applied Biosystems, USA) was used for reverse transcription of the validation cohort (n =6 [PE], 6 [NP]). Quantitative polymerase chain reaction was performed on Quant Studio^TM^ 3 real-time PCR systems, (Quant Studio^TM^ Design and analysis software), MicroAmp^TM^ Optical 96-well reaction plate (N8010560) and optical adhesive film kit (4313663), with the hydrolysis probe-based Taqman (R) gene expression assay (Applied Biosystems, USA). The following settings were used: hold 50°C for 2 minutes, hold at 95°C for 20 seconds, followed by forty 95°C for 1 second and 60°C for 20 seconds. Cq values were generated automatically by the Quant Studio Design and Analysis desktop software. The geometric mean of *YWHAZ* and *SDHA* were used as the internal reference genes for the normalization of all qPCR data. We analyzed the qPCR data and calculated fold changes using the 2^-ΔΔCt^ method (Livak & Schmittgen, 2001). Data was expressed as fold change, and standard error is denoted as error bars with GraphPad Prism software (version 9). Statistical testing was made on the ΔCt values using a one-tailed Student *t-test,* and significance was set at *P* < 0.05. All gene expression assays, their corresponding assay IDs, and all other details used in this study are listed in supplementary material.

## Results

### Patient demographics and clinical characteristics

There were no significant differences in maternal age, body mass index, and the gender of the neonates (Table 1). The average systolic (178.83 mmHg) and diastolic (109.17 mmHg) blood pressure was significantly higher among the PE cohort (P<0.001). Likewise, there was a significant difference in proteinuria (PE = 2.58; NP = 0 pluses on urine dipstick; P < 0.001) and gestational age at delivery (PE = 32.00 weeks gestation, NP = 39.17 weeks gestation P = 0.001) in PE compared to NP. Finally, PE neonates were more likely to be growth restricted (100%; 0% P = 0.004) with an average birth weight of 1515.83 g compared to 3912.50 g in normal neonates (P < 0.001).

**Table 1.**
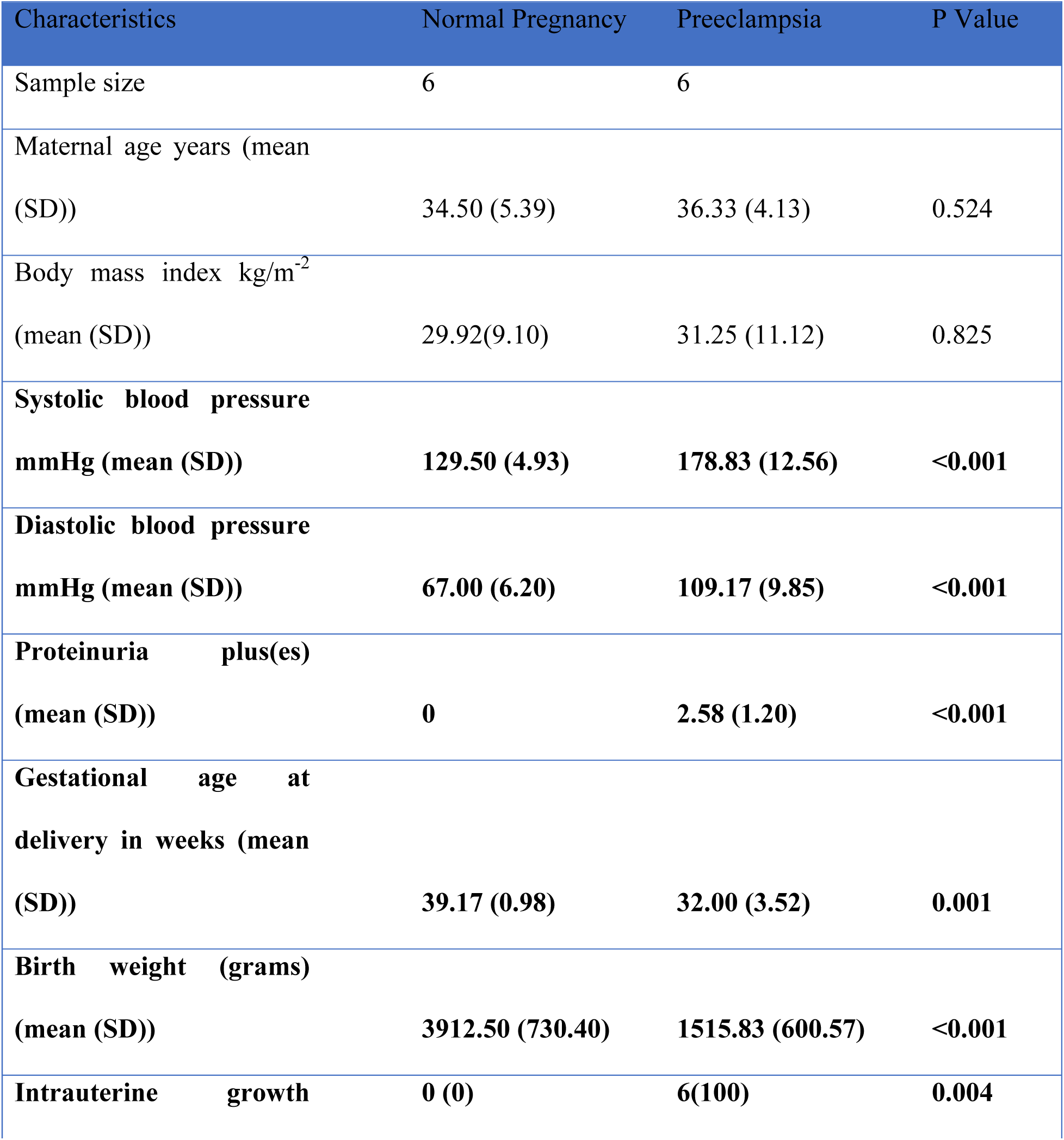
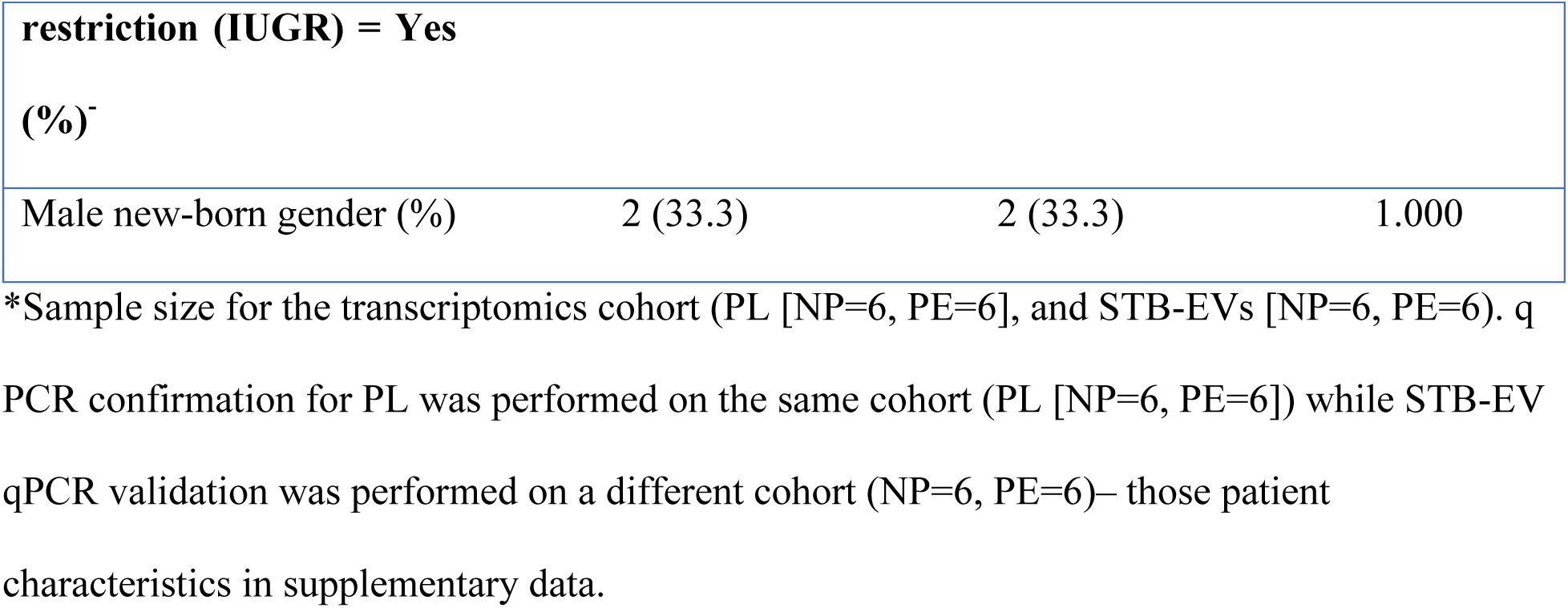
General characteristics of the transcriptomic cohort study population.

### Characterization of STB-EVs by TEM, NTA, flow cytometry, and WB

We characterized our sample preparations with transmission electron microscopy (TEM), nanotracking analysis (NTA), flow cytometry (FC), and western blot (WB) after the isolation and enrichment of STB-EVs. TEM (Figure 1A) showed the typical cup-shaped morphology of extracellular vesicles. In particular, the 10K STB-EV pellet (Figures 1A2, 1A5 and 1A8) showed a size heterogeneity characteristic of m/lSTB-EVs (221 to 1000 nm) and the 150K STB-EV pellet (Figures 1A3, 1A6, and 1A9) showed a homogeneous EVs size profile (less than or equal to 220 nm). This finding was also replicated by NTA (Figure 1C). The 10K STB-EV (Figure 1C1) pellets had a modal size of 479.4 ± 145.6nm, while the 150K STB-EV (Figure 1C2) pellets had a smaller modal size (205.8 ± 67.7nm). The post 10K and post 150K pellets are hereafter renamed m/lSTB-EVs and sSTB-EVs, respectively. WB (Figure 1B) detected PLAP (66 KDa), tetraspanins (CD63 [30–65 KDa]), and endosomal trafficking proteins (ALIX [95 KDa]). The non-EV marker cytochrome C (12 KDa) was detected in the placenta lysate but not the STB-EV fractions. Notably, TSG101 and ALIX were more prominent in the sSTB-EV fractions than the m/lSTB-EV fractions and placenta lysate.

**Figure 1.**
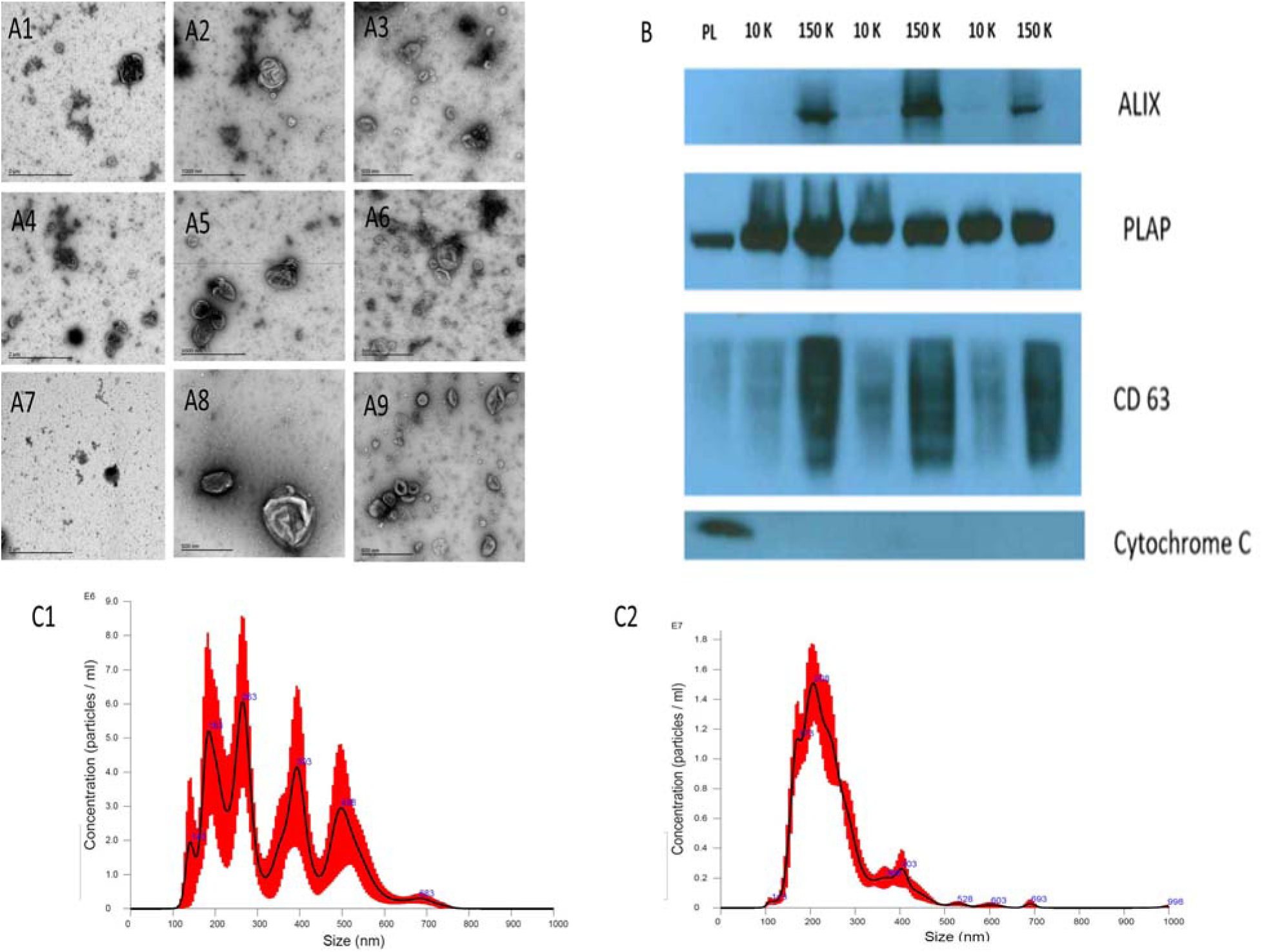
STB-EV characterization. Figure A displays representative transmission electron microscopy (TEM) images with wide view of samples (A1, A4, and A7), medium/large STB-EVs (A2, A5, and A8) displaying heterogeneity of vesicle sizes, and small STB-EVs (A3, A6, and A9) displaying typical cup shaped morphology. Figure 1B is a western blot of placenta homogenate (PL), m/lSTB-EVs (10K) and sSTB-EVs (150K) showing positivity for PLAP in all samples confirming placental origin. sSTB-EVs express enhanced levels of ALIX and CD63 which are known exosomal markers. The absence of cytochrome C in the STB-EV population confirms no contamination. Figure 1C shows the NTA results of m/l STB-EVs (C1) which show a broad size distribution and sSTB-EVs (C2) which display a more homogeneous size.

Flow cytometry (Figure S1) was only performed on the m/lSTB-EVs due to the size detection limit of flow cytometry which does not permit interrogation of small STB-EVs. We found 85 ± 8.3 % (Figure S1C) of qualified events were negative for the following lineage markers (CD41 [platelets] and CD235a [red blood cells], and HLA-I and II [white blood cells]). 95 ± 1.2 % of the EVs negative for non-placental markers (listed above) were BODIPY FL N-(2-aminoethyl)- maleimide (bioM) and placental alkaline phosphatase (PLAP) double-positive (Figure S1D). PLAP is a specific marker of syncytiotrophoblast, and bioM a marker of EVs, so this analysis confirmed that most of the post 10K samples, within the detection size range, were of placental origin. NP-40 (detergent) treatment confirmed that our samples were largely vesicular since only 0.1 ± 0.12%BODIPY FL N-(2-aminoethyl)-maleimide and PLAP double-positive events were detected (a reduction of 99%) after treatment with detergent (Figure S1E).

### Differentially carried genes (DCGs) in placenta homogenate, m/lSTB-EVs and sSTB-EVs between PE and NP

Transcriptomic analysis was performed on the discovery cohort (n=6 [PE], 6 [NP]). Comparison between PE and NP placental tissue revealed 580 upregulated and 563 downregulated (total) genes (adjusted P value of < 10^-5^ [Figures S3]) while in m/lSTB-EVs 1,128 were upregulated and 833 were downregulated (adjusted P value of < 10^-5^ [Figures 2]). In sSTB-EVs, 232 were upregulated, and 106 were downregulated (adjusted P value of < 10^-5^ [Figures S4]). We noted 25 downregulated genes, and 120 upregulated genes were common to all three sample types.

**Figure 2.**
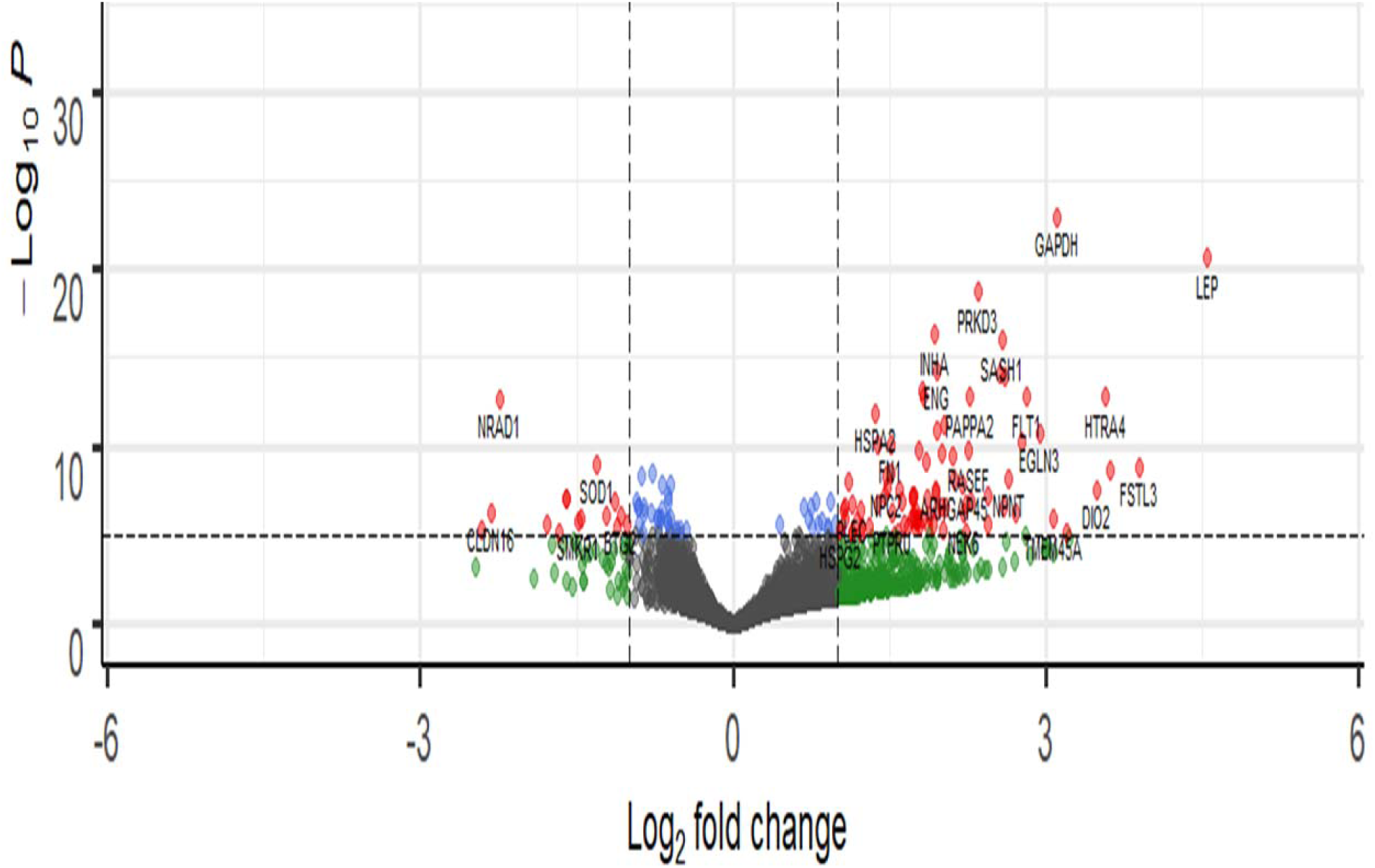
Representative volcano plot showing differentially expressed genes in medium/large STB-EVs. The most significantly upregulated genes are displayed in red on the right while the most significantly downregulated genes are displayed in red on the left. Volcano plots for placenta and small STV-EVs are in the supplemental data. -Log_10_P refers to the negative logarithm of the adjusted P Value.

### Quantitative PCR (qPCR) Validation of selected Genes

Quantitative PCR was performed on a different subset of STB-EVs, the validation cohort (n=6 [PE], 6 [NP]). As earlier discussed in the methods section, we identified the DCGs that were common between all three sample types (common DCGs) and of those, we selected a subset that were placenta enriched or specific based the criteria mentioned in the methods section. We selected *HTRA4, PAPP-A2, EBI3, HSD17B1, FSTL3, INHBA, SIGLEC6,* and *CGB3*. We performed qPCR analysis of these genes in both m/lSTB-EVs and sSTB-EVs. As a proof of concept, four of these genes known to be altered in PE in placental tissues (*LEP*, *COL17A1*, *SLC45A4*, and *FLNB)* were analyzed in the placenta and we confirmed that these genes were indeed significantly altered in PE (Figure 3). We next moved on to the species of interest and showed that in the m/l STB-EVs (Figures 3 and 4), *LEP*, *SIGLEC6*, *FLNB*, *COL17A1*, *SLC45A4*, *FSTL3*, and *HTRA4* were significantly different in PE compared to NP.

**Figure 3.**
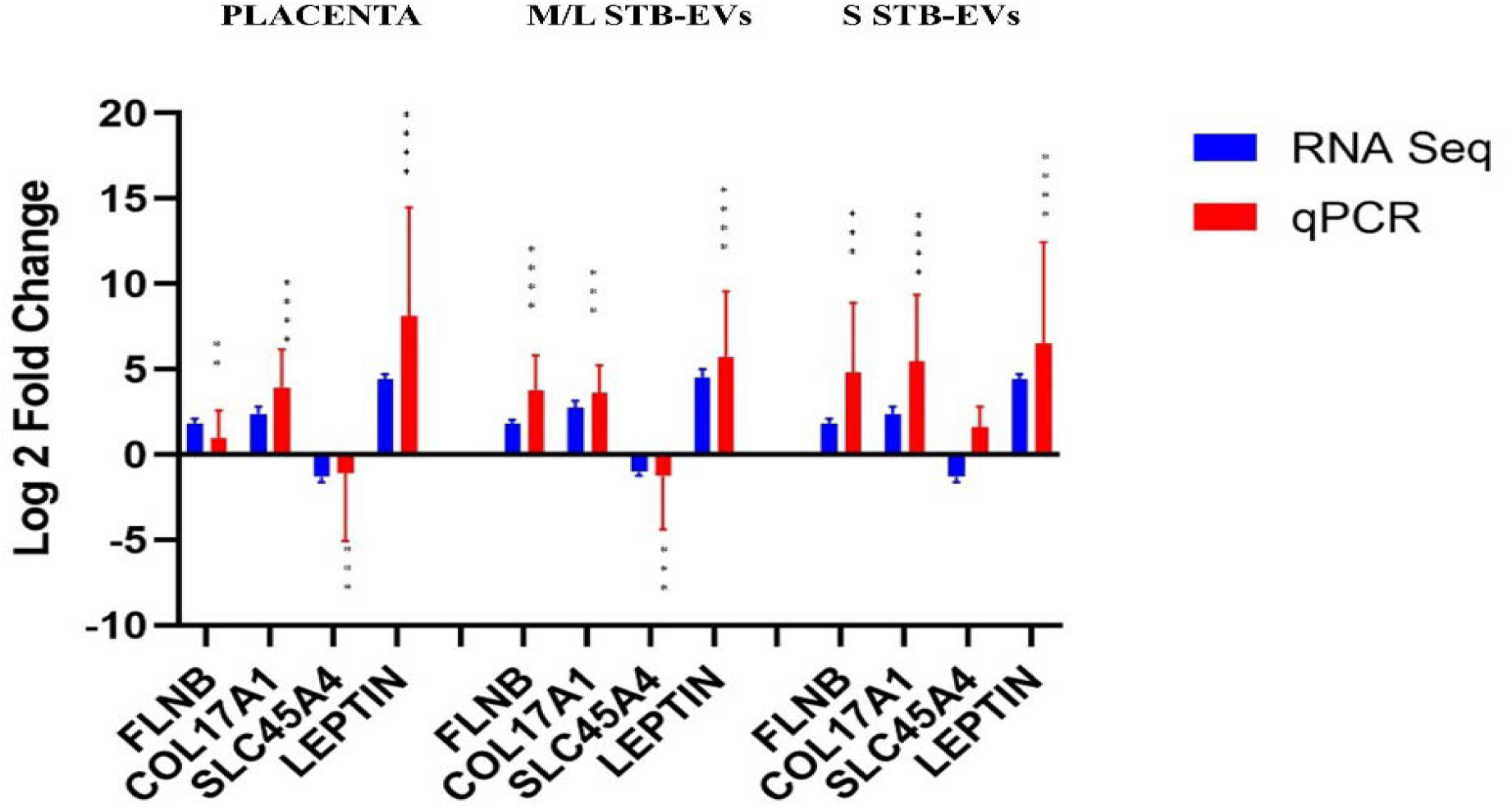
Quantitative PCR validation of selected differentially expressed/carried genes leptin (LEP), collagen type XVII alpha 1 chain (COL17A1), solute carrier family 45 member 4 (SLC45A4), filamin B (FLNB) and endoglin (ENG) in the placenta, medium/large STB-EVs and small STB-EVs. Data is visualized as mean fold change and error bars represent standard error. *=P <0.05, **=P <0.01, ***=P <0.001, and ****=P <0.0001. Sample size (n) = 6(NP) and 6(PE)

**Figure 4.**
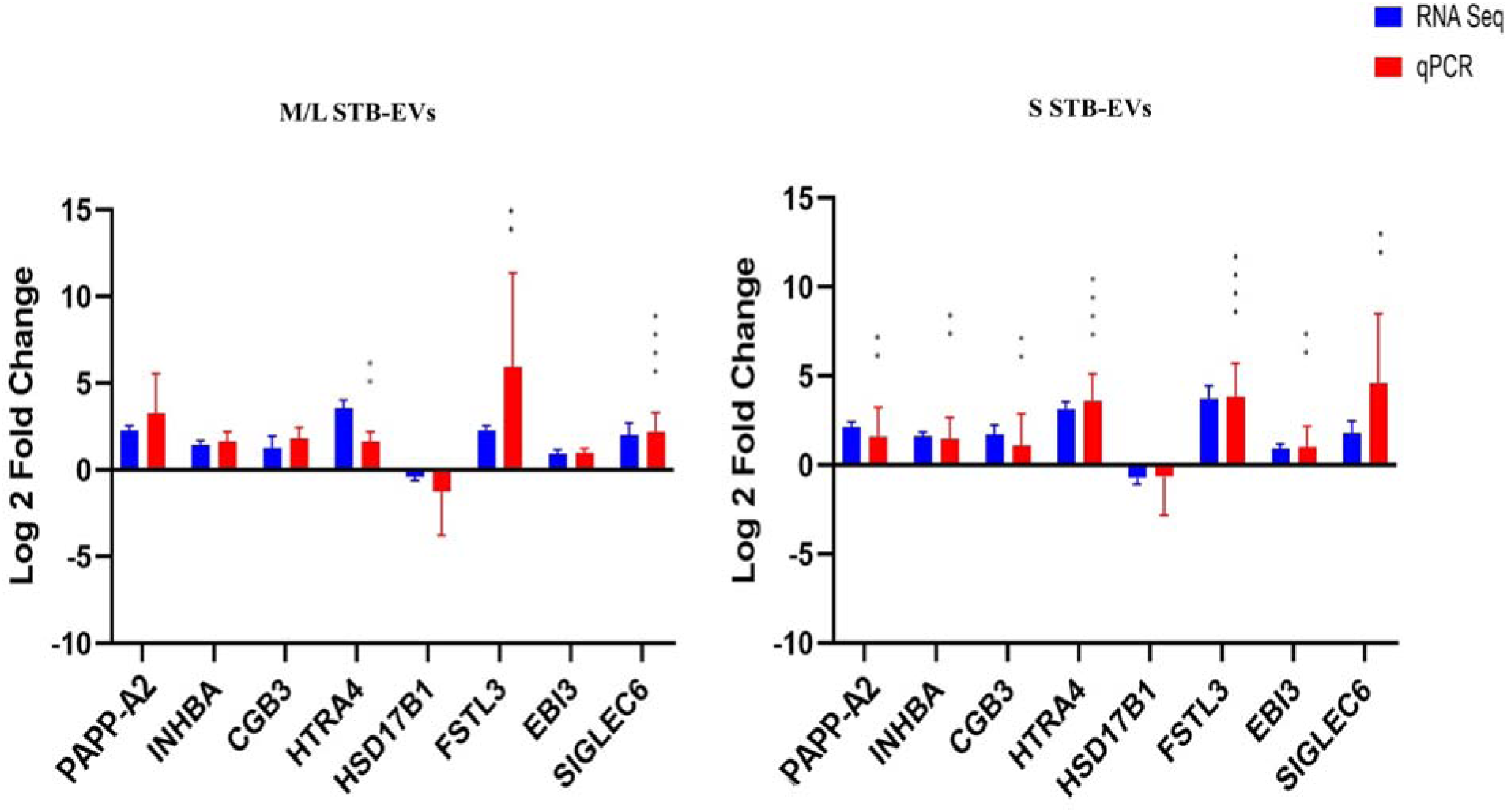
Quantitative PCR validation of placenta specific genes; pappalysin 2 (PAPP-A2), inhibin subunit beta A (INHBA), sialic acid binding Ig like lectin 6 (SIGLEC6), HtrA serine peptidase 4 (HTRA4), Epstein-Barr virus-induced 3 (EBI3), follistatin like 3 (FSTL3), hydroxysteroid 17-beta dehydrogenase 1 (HSD17B1), chorionic gonadotropin 3 (CGB3) and age-related macular degeneration (ARMS2) in medium/large STB-EVs and small STB-EVs based on qPCR. Data is visualized as mean fold change, and error bars represent standard error. *=P <0.05, **=P <0.01, ***=P <0.001, and ****=P <0.0001. Sample size (n) = 4(NP) and 4(PE).

In sSTB-EVs, (Figure 3 and 4) all the selected genes (except for *SLC45A4 and HSD17B1)* were significantly different. Across the three sample types, *LEP, COL17A1,* and *FLNB* were all significantly different between PE and NP while *SIGLEC6, FSTL3,* and *HTRA4* were significantly different in both m/l and sSTB-EVs.

### Functional enrichment analysis and Signalling pathway impact analysis (SPIA) of differentially expressed genes (DEGs) in preeclampsia (PE)

To understand the role of these genes and perhaps STB-EVs in the pathophysiology of preeclampsia, we performed a functional enrichment analysis (with overrepresentation analysis) on the list of DCGs (between NP and PE) for placenta tissue, m/lSTB-EVs, and sSTB-EVs and identified functional processes that overlap among the three sample subtypes. There were 32 similar biological processes between the placenta and sSTB-EVs (See full list Table S4). One biological process was similar between the placenta and m/lSTB-EVs (*platelet degranulation),* two biological processes; c*ell adhesion molecule* and *integrin-binding* common to both placenta and sSTB-EVs and no biological process was common to all sample subtypes.

When analysing KEGG pathways, *focal adhesion* was overrepresented among all three sample types while the *HIF-1 signalling pathway, proteoglycans in cancer, central carbon metabolism in cancer* were overrepresented in both placental tissue and sSTB-EVs.

Signalling pathway impact analysis of the DEGs in placental tissue homogenate showed *neuroactive ligand-receptor interaction, ECM-receptor interaction, focal adhesion, amoebiasis,* and *gap junction* as the most overrepresented. Of these five, all were inhibited except the *neuroactive ligand-receptor interaction,* which was activated. In contrast, the same analysis on m/lSTB-EVs revealed two significantly dysregulated pathways, *focal adhesion,* and *cytokine-cytokine interaction pathways,* both of which were activated. Similarly, in sSTB-EVs, three pathways *adipocytokine, focal adhesion,* and *type II DM* were significantly activated. Among all three sample types, *focal adhesion* was common to all.

## Discussion

### Principal findings

In recent years, extracellular vesicles (EVs) have gained interest, particularly their role in the context of PE, their potential to affect functional changes in organs distant from the placenta, and their ability to act as circulating liquid biopsies, revealing data about the donor cells in real-time. Our analysis found the STB-EV transcriptomic signature in PE to be different than in NP. We identified and validated STB-EV mRNAs—;FLNB, COL17A1, SLC45A4, LEP, HTRA4, PAPP-A2, EBI3, HSD17B1, FSTL3, INHBA, SIGLEC6, and CGB3. Exploration of these differentially abundant mRNAs as circulating biomarkers may help early identification of PE.

### Results in the context of what is known

Some of the findings of this study such as *FLT1*, *LEP* (Tsai et al., 2011b), *ENG*, *PAPP-A2* (Guo et al., 2021), *FSTL3, PRG2* (Gormley et al., 2017), *INHBA* (Brew et al., 2016), have been well described in the placenta but not previously described in STB-EVs. These STB-EV associated mRNA may account for some circulating mRNA such as *PAPP-A2*, *LEP*, and *HTRA4* (Munchel et al., 2020) detected in increased amounts in PE patients. We opted to validate the following genes: *FLNB, COL17A1, SLC45A4, LEP, ENG*, *HTRA4, PAPP-A2, EBI3, HSD17B1, ARMS2, FSTL3, INHBA, SIGLEC6,* and *CGB3* since these fulfilled the criteria for biomarkers as outlined.

We discovered that on m/lSTB-EVs, *FLNB, COL17A1, SLC45A4, LEP, FSTL3, HTRA4* and SIGLEC*6* were significantly differentially expressed between PE and normal. Equally interestingly, in sSTB-EVs we found that except for HSD17B1, all the other genes were differentially carried between PE and normal.

LEP was the most significantly elevated gene in placental tissue, m/lSTB-EV and sSTB-EVs. The discovery of LEP in STB-EVs is novel. The role of its protein, not its mRNA, as a potential biomarker has been suggested previously(Taylor et al., 2015). LEP plays a role in endocrine functions, reproduction, immune function and is produced by the placenta, aiding implantation, and regulating placental growth (Castellucci et al., 2000). The dysregulation of leptin has been linked to pregnancy complications with higher circulating protein levels associated with PE (Masuyama et al., 2010).

Similarly, we found significantly elevated COL17A1 in the placenta, m/lSTB-EV and sSTB-EV. COL17A1 is a collagen transmembrane protein involved in extracellular matrix receptor interactions. It has been identified, through mass spectrometry, to be differentially expressed in urine samples from PE. In contrast, we found that SLC45A4 mRNA was significantly downregulated in both types of vesicles. *SLC45A4* protein is involved in promoting glycolysis and autophagy prevention (Chen et al., 2021), two processes found to be abnormal in PE (Bloxam et al., 1987).

*FLNB* is an essential cytoskeletal protein critical in endothelial cell motility and that has been previously reported to be downregulated in PE placentae at the mRNA and protein level(Tejera et al., 2013; Wei et al., 2019). This contrasts with our study which showed that FLNB gene was significantly elevated in placenta and in m/lSTB-EV and sSTB-EV. It is difficult to identify the exact reason for the variance, but the choice of housekeeping gene may be a possible explanation. Wei et al used *β actin* which has been observed to be variable and a potential confounder in other disease states or experimental conditions (Glare et al., 2002). We used the geometric mean of *YWHAZ* and *SDHA*; both previously shown to be stably expressed(Meller et al., 2005)

Our analysis also showed that SIGLEC6 was differentially present in both subset of STB-EVs. Our group had previously reported the presence in higher concentration in PE placentas compared to NP(Awoyemi et al., 2020). *SIGLEC6* regulates immune cell by interacting with sialic acid on these cells and inhibiting cellular activation and clonal expansion (Zhang et al., 2007) through cytosolic tyrosine-based regulatory motif (Brinkman-Van der Linden et al., 2007). *FSTL3* glycoprotein is localized to the decidua and has been well documented to be present in abundance in PE placenta and plasma samples. (Xie et al., 2018), (Founds et al., 2015). Our analysis demonstrates that this elevation is also mirrored at the mRNA level in STB-EVs. *HTRA4* was found to be differentially carried in PE STB-EVs in our analysis. HTRA4 is a placental-specific serine protease that inhibits cell cycle progression and cell differentiation to endothelial cells (Wang et al., 2019). *HTRA4* may be responsible for endothelial cell dysregulation by disrupting endothelial tube formation and increasing endothelial cell permeability (Singh et al., 2015).

We also attempted to provide mechanistic insights into the roles of these differentially carried genes in the pathogenesis of PE. We identified *HIF 1 pathway, blood vessel development* (Guo et al., 2021)*, adipocyte regulation, retinoic acid regulation, hypoxic signaling, associated cytokine pathways* (Benny et al., 2020)*, PI3/Akt signaling,* and *pathways in cancer* (Li & Fang, 2019) as pathways that are potentially contributory. We also found several less well characterized and potentially relevant pathways such as *processes involved with protein catabolism and degradation, insulin and other growth factor receptor signaling pathways, nervous system development, ion channel activities, transcription repression,* among others. Interestingly, the signalling pathway impact analysis found that the *focal adhesion pathway* was inhibited in the placenta but activated in both STB-EV subtypes. C*ytokine-cytokine interactions* were activated in m/lSTB-EVs, while *type II DM* (diabetes mellitus) and *adipocytokine* pathways were activated in sSTB-EVs. The risk of PE is known to be increased by 2-4-fold among type II DM patients, and women with a history of PE are more likely to develop type II DM later in life(Weissgerber & Mudd, 2015). None of the patients used in this study had been diagnosed with diabetes mellitus. If they had, it may have explained why this pathway was activated in our analysis. It may be interesting to explore, in the future, if sSTB-EVs play a role in this association or if this association is due to the presence of comorbid conditions common to both PE and DM.

Finally, *focal adhesion* is a critical step in cell and extracellular matrix (ECM) interaction and perhaps interactions between the trophoblast and the ECM and is crucial in trophoblastic invasion and PE (He et al., 2015). In our analysis, we found this pathway to be inhibited in the placenta but activated in m/l and sSTB-EVs. It is reasonable to assume that the differences in the GO terms and pathways among all sample types may portray a difference in roles between the placenta, M/L STB-EVs, and S STB-EVs in the pathogenesis of the disease. However, we believe these differences may be best explored in futured studies.

### Ideas and Speculations

Our findings have identified several potential STB-EV mRNA biomarkers which require external validation especially in biofluids like plasma, serum, or urine. The prospect of a qPCR biomarker panel is interesting as a qPCR panel should be cheaper and easier to implement than a quantitative protein assay. Such tests are easily scalable and if validated might even be adaptable to low-income resource settings using Loop-Mediated Isothermal Amplification (LAMP) PCR, a PCR technique than can be performed at room temperature. Since transcriptome level alterations precedes proteome level alterations, mRNA based biomarkers may theoretically be better at ascertaining the risk of developing preeclampsia before the development of clinical symptoms than standard of care.

### Strengths and limitations

Our study is not without limitations that should be considered while interpreting the results. First, our sample size is relatively small and thus no predictive analysis could be conducted. Secondly, it is difficult to find a gestational age matched control for PE, particularly the early onset phenotype (< 34 weeks). Therefore, we used term (37 to 41^+^ ^6^ weeks) controls. This is a standard approach for all scientists working on PE placentas. Lastly, like most studies on PE, the samples are from deliveries. It is possible these biomarkers represent differences observed at end-stage disease and would require further research to translate these markers to early developmental stages. Notwithstanding, this study has numerous positives, particularly our ability to utilize a comprehensive analysis of three linked sample types from the same patient. This has permitted the dissection of differences between the tissue and the EVs and identification of some potentially exciting pathways which may form the basis of further research.

## Conclusions

Despite intensive efforts, the pathophysiology of preeclampsia (PE) has still not been completely unraveled. In this study, we identified potential mRNA biomarkers and mechanistic gene pathways that may be important in the pathophysiology of PE and could be further explored in future studies. The potential utilization of STB-EVs based mRNA as circulating biomarkers with real time placental and fetal information may be important in early PE diagnosis and mechanism.

## Data Availability

All data produced are available online at DOI: 10.17632/6s5fkd3z7t.1

https://dx.doi.org/10.17632/6s5fkd3z7t.1

## Acknowledgments

We acknowledge the support of the National Institute of Health Research Clinical Research Network Fenella Roseman & Lotoyah Carty research for assistance in patient recruitment.

## Supplemental Material

### Supplemental Methods

#### Enrichment of STB-EVs by placental dual-lobe perfusion and serial ultracentrifugation

Briefly, we identified a suitable cotyledon (devoid of calcifications, ischemia, or rupture) and cannulated a placental artery and vein perfusing the placenta for three hours at a 4-5 ml/min flow rate to obtain placenta perfusate. The placenta perfusate was centrifuged twice at 1,500 g for ten minutes at 4°C (*Beckman Coulter Avanti J-20XP centrifuge using a Beckman Coulter JS-5.3 swing-out rotor*) to remove cell debris. The supernatant was carefully pooled and spun at 10,000 g (10K) in a swing bucket centrifuge (*Beckman L80 ultracentrifuge and Sorvall TST28.39 swing-out rotor*) at 4^0^C for 30 minutes. The 10K STB-EV pellet was washed with filtered phosphate buffer saline (fPBS) followed by resuspension of the 10K STB-EV pellets in fPBS. An aliquot of the resuspended pellets was analysed to identify and characterise STB-EVs, while the rest were aliquoted to obtain a protein concentration around 2-5 µg/µl (measured using a Pierce bicinchoninic acid (BCA) protein assay) and immediately stored at -80°C. The post-10K supernatant was filtered through a 0.22 µm Millipore stericup filtration device, then spun at 150,000 g for 2 hours (*Beckman L80 ultracentrifuge with a Sorvall TST28.39 swing-out rotor*) and the 150K STB-EV pellets were washed, resuspended in fPBS and aliquoted like the 10K STB-EV pellets. This working stock was used for subsequent analysis.

#### Flow Cytometry

**Figure S1.**
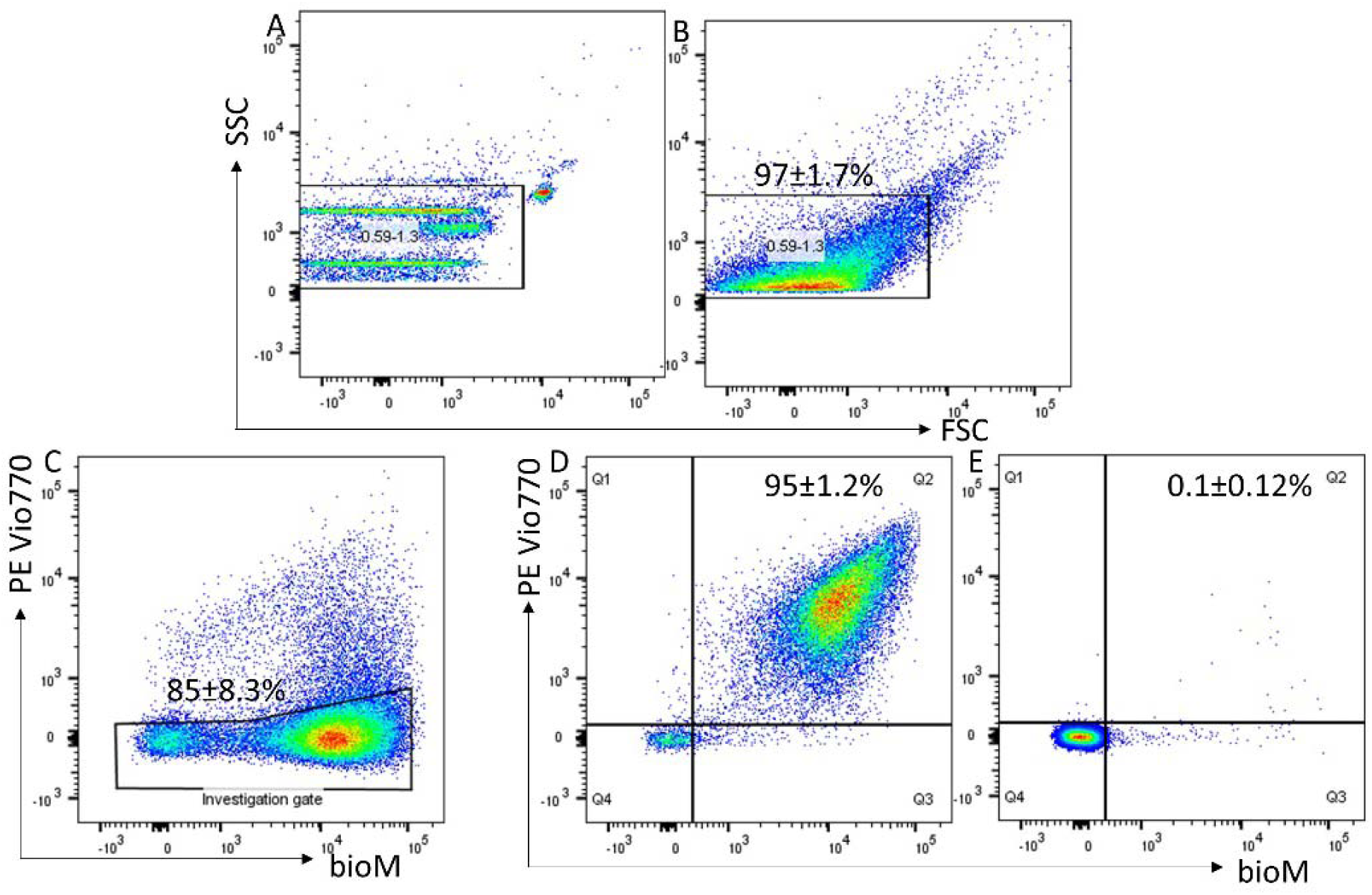
Representative flow analysis of m/lSTB-EVs enriched by placental perfusion. Apogee beads mix was used to set the flow machine’s light scatter resolution to 0.59-1.3 mm silica beads (Figure S1A); Application of SSC and FSC PMTVs determined by apogee beads mix for the analysis of M/L STB-EVs (Figure S1B). An investigation gate interrogating m/l STB-EVs which did not express the non-placental markers HLA Class 1 & II, CD41a and CD235a – thus removing co-isolated non-placental EVs (Figure S1C). STB-EVs from the investigation gate were further analyzed for expression of PLAP and staining by bioM-which stains proteins (Figure S1D). This revealed a high number (95%) of PLAP positive vesicles. These PLAP+ bioM+ double positive EVs were highly sensitive to detergent treatment (Panel E) with the reduction in PLAP+ bioM+ double positivity confirming that they are vesicular in nature.

**Figure S2.**
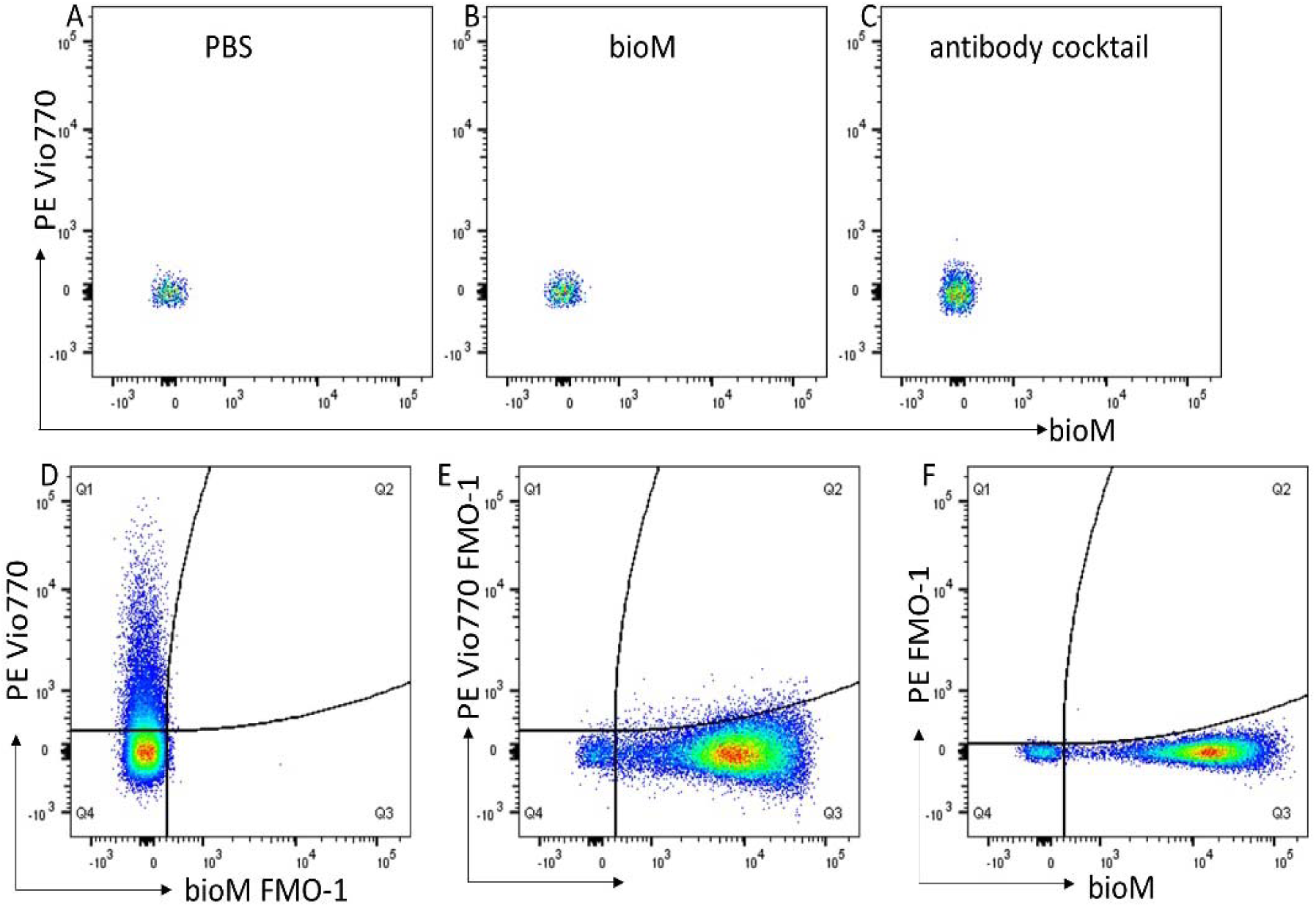
Reagent controls, 0.1mm filtered PBS (A); 0.2mm filtered bioM (B) and antibody cocktail (C) all showing no PLAP+ bioM+ double positivity. Fluorescent minus one (FMO-1) controls, bioM-1 control (D); PE Vio770-1 control (E); PE-1 control (F).

#### Western Blot Analysis

**Table S1.**
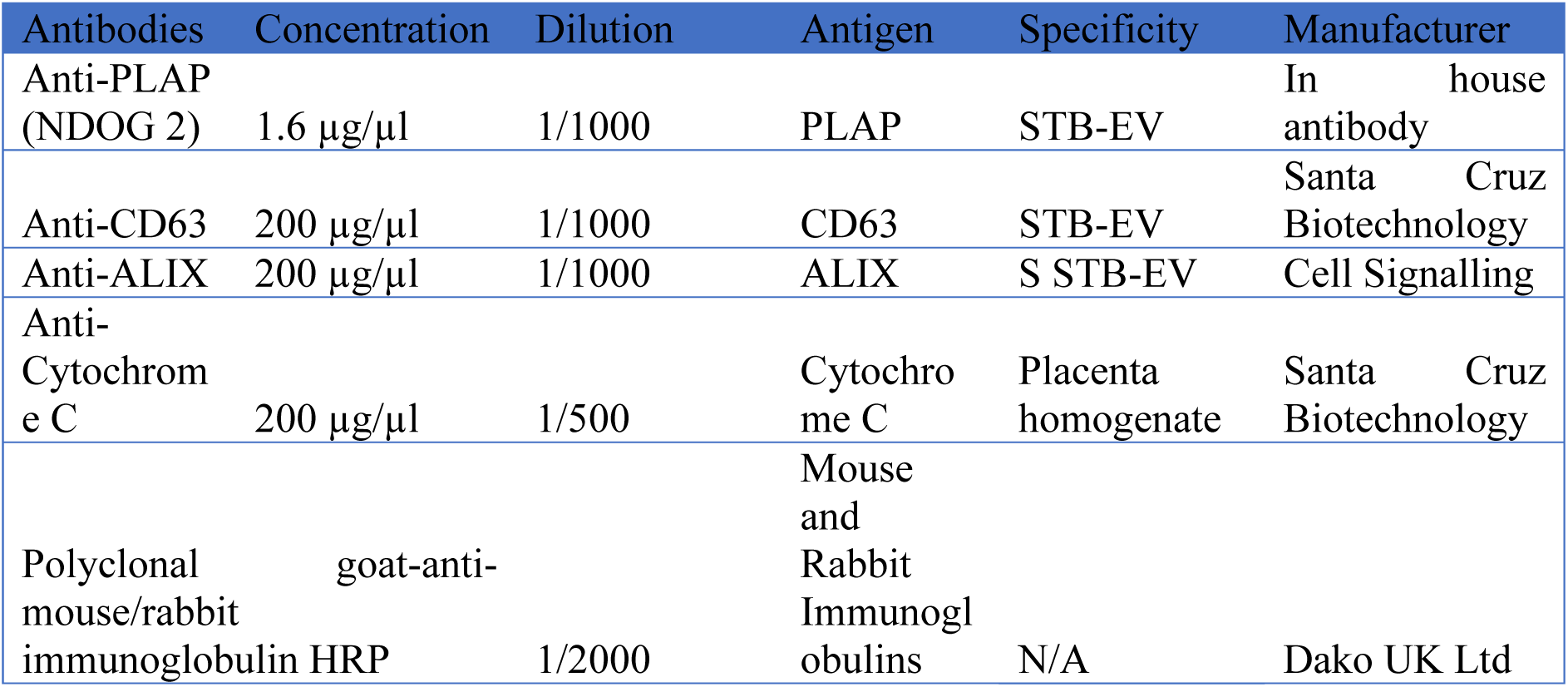
Table showing the antibodies used for Western blot.

#### RNA-Sequencing Library Preparation and Sequencing

For placental biopsy, 30 milligrams of placenta tissue were mechanically homogenized, and the RNA extracted with the RNeasy kit. An equal amount of STB-EV (200 µg) was diluted up to 200 µl. Total RNA was then extracted from this STB-EV mixture with the miRCURY™ RNA isolation kit. The extracted RNA was assessed for purity with Nanodrop (NanoDrop 1000 Software). After confirming that all samples had a 260/280 ratio greater than 1.8, the samples were sent to the Wellcome Centre for Human Genetics (WCHG) for sequencing. At WCHG, the RNA was assessed with a Bioanalyzer (Agilent Technologies) to determine the concentration and RIN values (for PL) which were all greater than 8 except the STB-EV samples with RIN between 1-3. It is widely accepted that the RIN values are not applicable for extracellular vesicles due to 1) the fragmented nature of RNA they contain and the limited presence of 18S and 28S ribosomal RNA peaks.

RNA libraries were prepared using the TruSeq RNA library preparation and small RNA library preparation kits (Illumina). A standard procedure was followed for mRNA sequencing. Briefly, polyadenylated mRNAs were selected from total RNA samples using oligo-dT-conjugated magnetic beads. Poly-adenylated RNA samples were immediately converted into stranded Illumina sequencing libraries using 200 bp fragmentation and sequential adapter following the manufacturer’s specifications. The resulting cDNA was amplified, enriched, and indexed using 12 cycles of amplification with PCR primers, including an index sequence to allow for multiplexing. All RNA sequences were purified on gels and sequenced on a HiSeq2500 high output v3 flow cell using paired-end, 75 bp reads (Illumina).

#### Taqman Gene expression assays

**Table S2.**
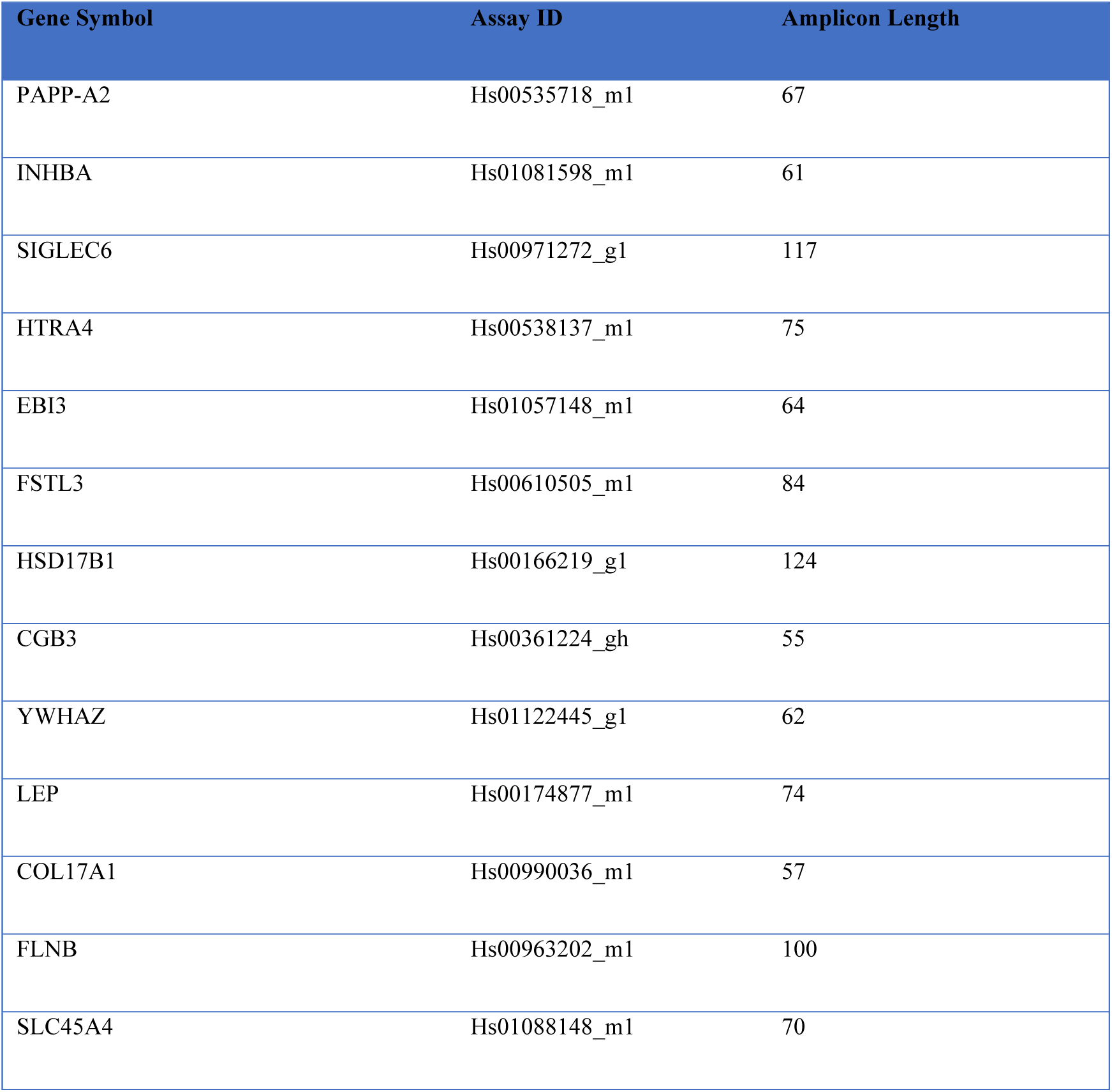
Table showing qPCR Primers, assay ID and amplicon length.

| Gene Symbol | Assay ID | Amplicon Length |
| --- | --- | --- |
| PAPP-A2 | Hs00535718_m1 | 67 |
| INHBA | Hs01081598_m1 | 61 |
| SIGLEC6 | Hs00971272_g1 | 117 |
| HTRA4 | Hs00538137_m1 | 75 |
| EBI3 | Hs01057148_m1 | 64 |
| FSTL3 | Hs00610505_m1 | 84 |
| HSD17B1 | Hs00166219_g1 | 124 |
| CGB3 | Hs00361224_gh | 55 |
| YWHAZ | Hs01122445_g1 | 62 |
| LEP | Hs00174877_m1 | 74 |
| COL17A1 | Hs00990036_m1 | 57 |
| FLNB | Hs00963202_m1 | 100 |
| SLC45A4 | Hs01088148_m1 | 70 |

### Supplementary results

**Table S3.** General characteristics of the STB-EV qPCR validation study population.

| Characteristics | Normal Pregnancy | Preeclampsia | P Value |
| --- | --- | --- | --- |
| Sample size | 6 | 6 |  |
| Maternal age years (mean (SD)) | 35.50 (4.80) | 37.25 (4.11) | 0.600 |
| Body mass index kg/m<sup>2</sup> (mean (SD)) | 24.25(3.30) | 34.88 (12.28) | 0.146 |
| **Systolic blood pressure mmHg (mean (SD))** | **109.50 (6.86)** | **183.75 (12.87)** | **<0.001** |
| **Diastolic blood pressure mmHg (mean (SD))** | **76.00 (1.41)** | **113.25 (9.71)** | **<0.001** |
| **Proteinuria plus(es) (mean (SD))** | **0** | **3.00 (0.82)** | **<0.001** |
| **Gestational age at delivery in weeks (mean (SD))** | **39.50 (0.58)** | **33.50 (3.00)** | **0.008** |
| **Birth weight (grams) (mean (SD))** | **3627.50(247.98)** | **1808.75(488.54)** | **0.001** |
| **Intrauterine growth restriction (IUGR) = Yes (%)** | **0 (0)** | **4(100.00)** | **0.034** |
| Male new-born gender (%) | 2 (50.00) | 1 (25.00) | 1.000 |

**Figure S3.**
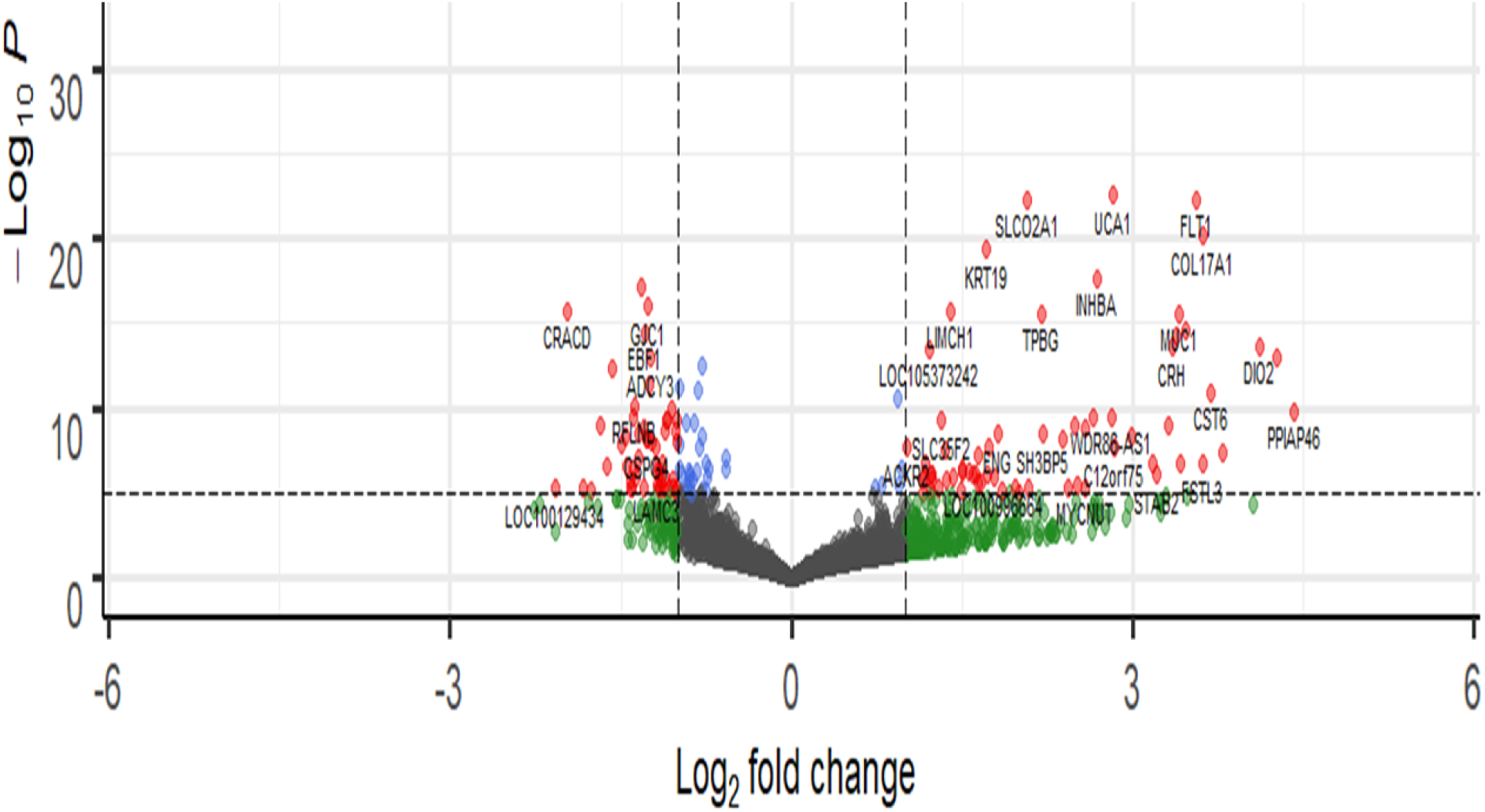
Volcano plot showing differentially expressed genes in the placenta. The most significantly upregulated genes are displayed in red on the right while the most significantly downregulated genes are displayed in red on the left.

**Figure S4.**
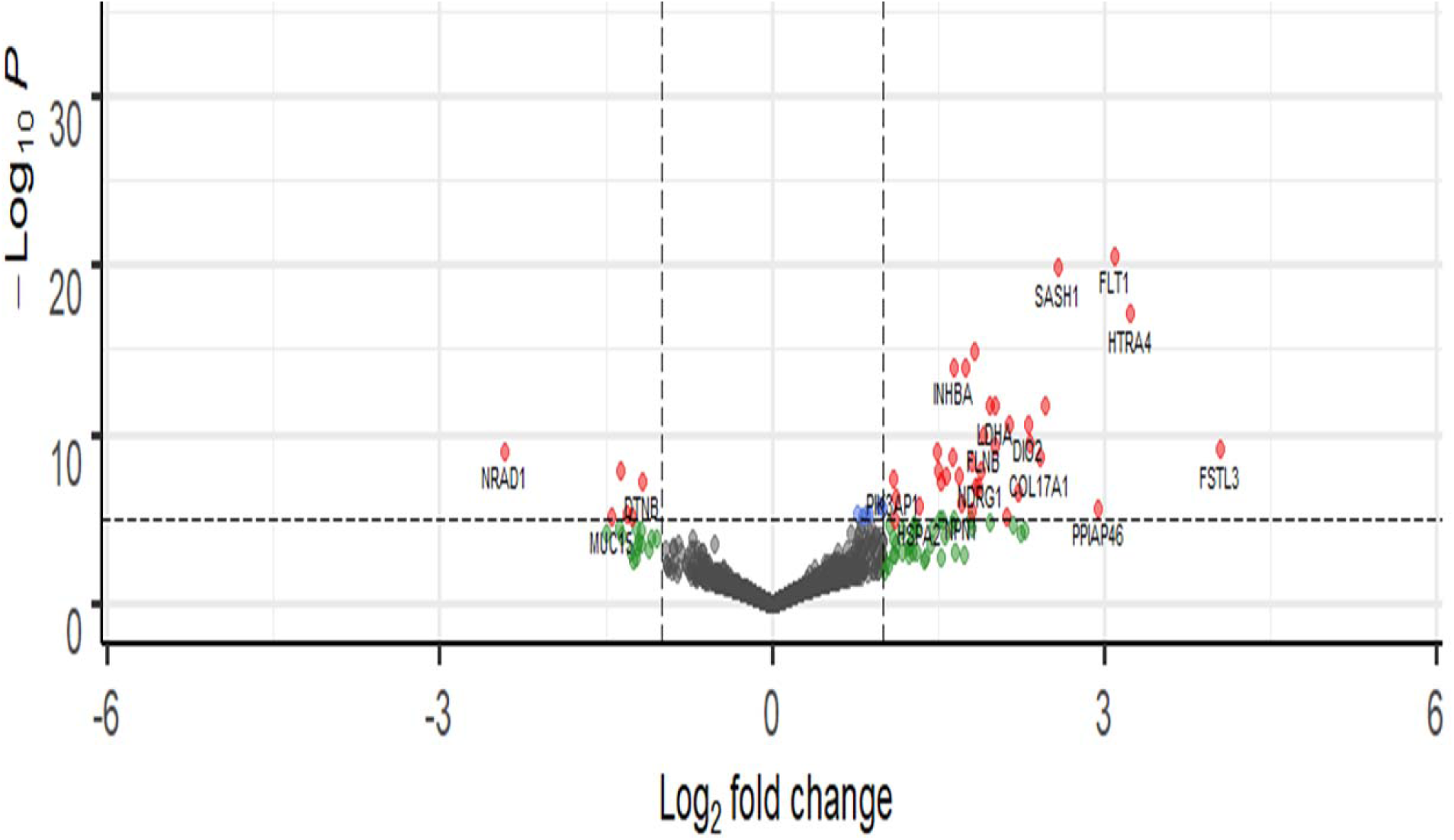
Volcano plot showing differentially carried genes in small STB-EVs. The most significantly upregulated genes are displayed in red on the right while the most significantly downregulated genes are displayed in red on the left.

**Table S4.**
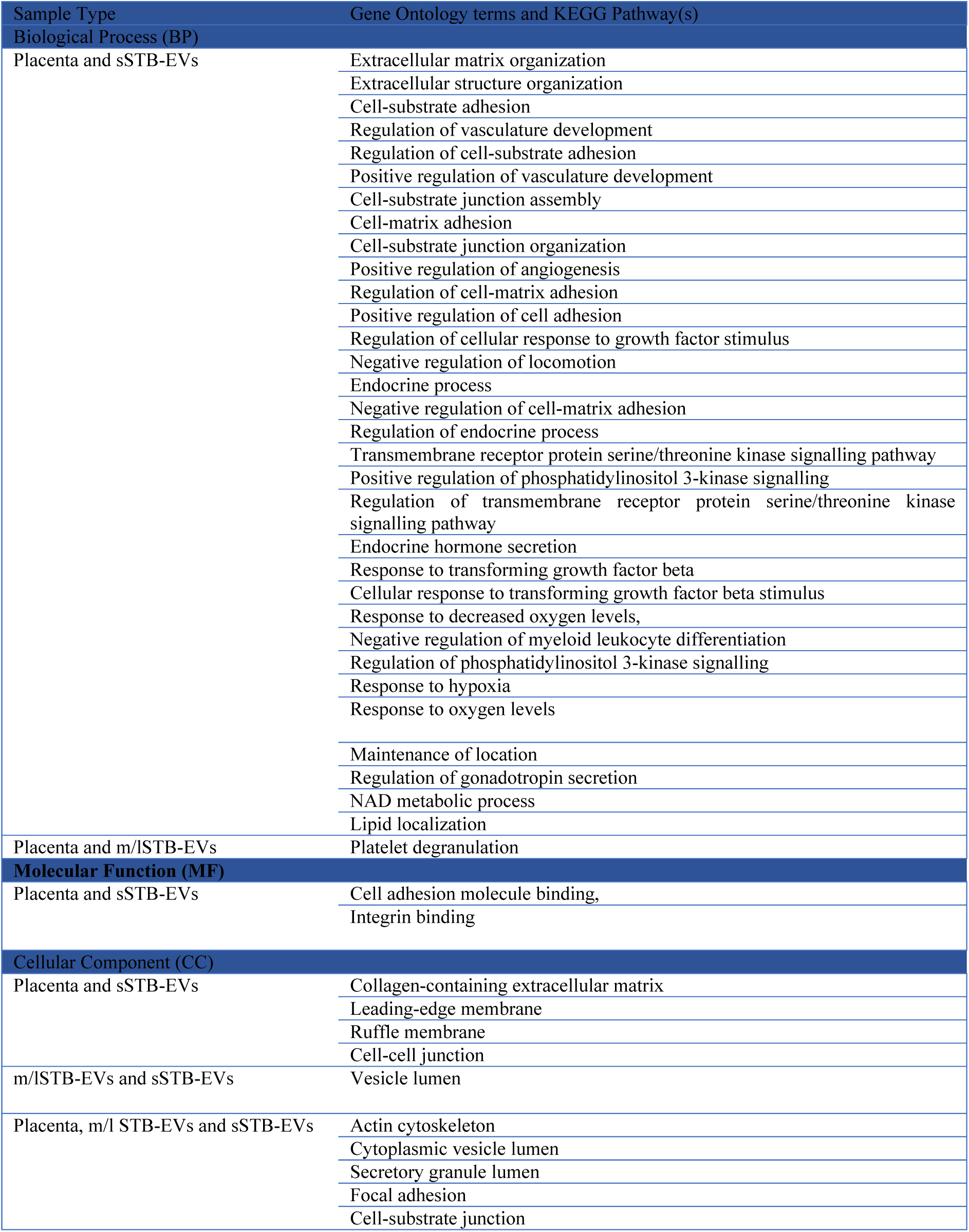
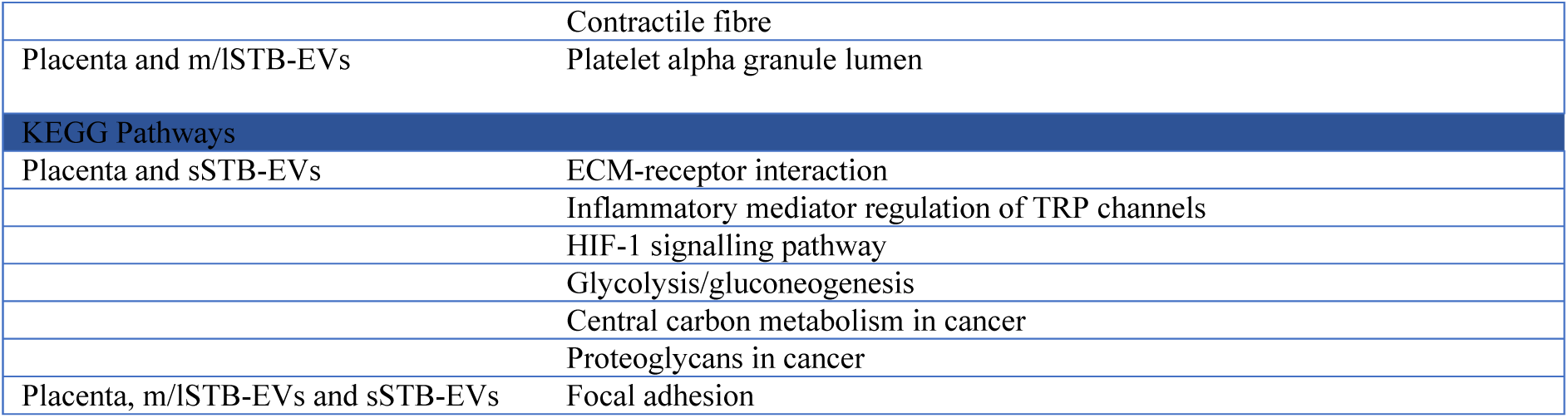
List of overlapping gene ontology (GO) terms and KEGG Pathways in the placenta, medium/large STB-EVs and small STB-EVs.

